# Studying the effect of lockdown using epidemiological modelling of COVID-19 and a quantum computational approach using the Ising spin interaction

**DOI:** 10.1101/2020.06.06.20124024

**Authors:** Anshuman Padhi, Sudev Pradhan, Pragna Paramita Sahu, S Kalyani, Bikash K. Behera, Prasanta K. Panigrahi

## Abstract

COVID-19 is a respiratory tract infection that can range from being mild to fatal. In India, the countrywide lockdown has been imposed since 24th march, 2020, and has got multiple extensions with different guidelines for each phase. Among various models of epidemiology, we use the SIR(D) model to analyze the extent to which this multi-phased lockdown has been active in ‘flattening the curve’ and lower the threat. Analyzing the effect of lockdown on the infection may give us a better insight into the evolution of epidemic while implementing the quarantine procedures as well as improving the healthcare facilities. For accurate modelling, incorporating various parameters along with sophisticated computational facilities, are required. Parallel to SIRD modelling, we tend to compare it with the Ising model and derive a quantum circuit that incorporates the rate of infection and rate of recovery, etc as its parameters. The probabilistic plots obtained from the circuit qualitatively resemble the shape of the curve for the spread of Coronavirus. We also demonstrate how the curve flattens when the lockdown is imposed. This kind of quantum computational approach can be useful in reducing space and time complexities of a huge amount of information related to the epidemic.

## I. INTRODUCTION

COVID-19 (Coronavirus Disease 2019) is a disease caused by the virus strain known as SARS-CoV-2 (Severe Acute Respiratory Syndrome Coronavirus II). It has widespread implications on the human body in the form of respiratory issues, septic shock, co-morbidity arising from multiple-organ failure, and even death [1]. Routing back to its emergence in mainland China around the end of 2019, till May 2020, it has spread to over 210 countries resulting in a total of around 40 lakh cases with almost 3 lakh deaths due to the same. The World Health Organisation (WHO) declared it as a global pandemic on 11th March 2020, observing the rate it transmits. Various countries, including India, put forward extensive measures to curb the viral epidemic, by extensive tracing, testing and isolating the suspected ones while improving healthcare systems and imposing lockdowns. Govt. of India declared the countrywide lockdown on 24th March 2020 to reduce the virus’s rate of transmission. To tackle this global pandemic, the extent of spread and the time taken by the epidemic to reach its peak and other details must be well predicted so that the state can plan accordingly and fight against it.

Mathematical modeling can come handy in these processes, as they can predict how the epidemics evolve while analyzing the current set of available data. Any such prediction system requires a set of assumptions and considerations, which helps to formulate the necessary equations that can later project the regular convolutions. Precise consideration of networks within a population while the model is being formulated yields an accurate prediction. Later, the prediction can be informed to the healthcare sector and the stakeholders for necessary implementations.

Here, we use a time-dependent SIRD (Susceptible-Infected-Recovered-Deceased) model to predict the evolution of this epidemic in India. The SIRD model is one among several compartmental studies in epidemiology [2] such as SEIR, where E stands for exposed and rest staying the same; SEIRD, SIR, SIS model, etc. They have its origin from the Kermack-McKendrick theory of infection spread, a very rigorous statistical analysis performed in 1927. Here, the population is divided into various compartments, and their interactions are studied further. It is a simple yet an instrumental model of epidemiology since it takes into account various factors such as the rate at which infection spreads, the rate at which the active cases recover, etc. In this current study, we use the data present at the crowd-sourced COVID-19 tracker [32]. We use classical computation to demonstrate how a change in spread rates might stabilize figures related to infection, recovery, and death.

Several studies have been done with the available set of data of COVID-19 spread in India and other countries. Some of the notable works have used several other statistical approaches while some have used compartmental studies to model and predict the viral spread [4–12].

In our study, we focus on breaking the time scale into smaller intervals (a period of ten days) and analyzing them individually to determine the value of the parameters specified in these periods. We demonstrate how step-wise constraints on such systems have effectively reduced infection spread over time. Here, we introduce two possibilities concerning a set of parameters, one which depicts a situation with no lockdowns and another with controlled movement of people. We discuss and analyze the curves obtained from the simulation in terms of rates of infection spread, as they are mostly dependent on the interaction network of subjects in a concerned system explicitly. As we study this parameter, we aim to establish a pattern in their evolution based on data obtained and extrapolate them to obtain a specific value in the timeline towards the end of the multi-phased lockdown (supposedly on 17th May). Further, this set of predicted parameters is used to deduce the progress of the system in the near future with a fixed population. These deductions with current constraints on the system are put up with a system with no such constraints to demonstrate the efficacy of multi-phased lockdown in widening and delaying the peak of infection reporting.

Quantum computation can be useful in the assessment of such systems since an accurate prediction of a viral spread needs to encompass various factors that might pose complexity challenges in classical computation. Factors like quarantine measures, social distancing, population networking, self-protection actions, etc, can give rise to a complex set of problems, challenging for a classical computer to solve. Assuming such factors could be easily fitted into quantum computation facilities, given its intrinsic ability to hold substantial information and parallelly process them, underlies this project’s hypothesis.

Parallel to the SIRD modelling in the study, we form an analogy to the Ising model of magnetic lattice to form a Hamiltonian. Then we build a quantum circuit to demonstrate how they are efficient enough to qualitatively show the nature of the epidemic through the obtained graphs as outputs. We also demonstrate how the considered parameters must be varied to reduce the number of infections when at its peak and also to delay the time by which viral spread peaks in the country. This delay of the peak with lowering of its height is given a term called ‘flattening the curve’ and this becomes crucial, as the affected population is more spread out for a given time interval. Hence, this might not flood the country’s healthcare facilities, unlike otherwise. The reduction of the number of cases at its peak also ensures that the current healthcare facility faces a shortage of resources while treating the patients.

To build the quantum circuit, we have used the IBM Quantum Experience platform. Various prototypes of quantum operators have been designed and have been made available through IBM quantum experience, a free-web based platform. Researchers have used it to their strength to experiment with circuits and also to simulate results which have furthered their research [16–31].

We organize this paper as follows. Section II discusses the theoretical backgrounds of the SIRD compartmental model and the quantum computational approach. Section III proposes how both of them have been implemented and what results were obtained from them. Section IV is dedicated to a general discussion on the obtained results, including the drawbacks. Furthermore, at last, we conclude this article by citing the future implications of the proposal in Section V. For additional references, an appendix has been attached at VI.

## II. THEORETICAL BACKGROUND

### A. SIRD Model

As already mentioned, the SIRD model is one of the compartmental models used in epidemiology. It divides the whole population into categories where S stands for the part of the population which is susceptible to being infected by the virus. I is the population that has been infected and has the potential to spread the infection. R is the group which has successfully recovered from the disease. D stands for the portion which has been deceased after getting infected due to it. N, the country’s total population assumed to be time-independent, is the sum of the susceptible, infected, recovered and the deceased.

To develop the operating mathematical model equations, some assumptions have been made here to keep the model computationally simple. They are

- The average birth rate and mortality rate of India have not been considered.
- The mode of transmission has been considered to be from person to person.
- Once a person has recovered from the disease, he/she has attained the immunity for infection, hence does not fall back to the category of susceptibles.

Here *α* is the rate at which the infection is transmitted to the susceptibles due to possible contact between infected and susceptible ones. *β* indicates the rate at which the infected individuals recover, which is the reciprocal of the number of days in the treatment period. *γ* describes the fatality of the virus as the rate at which infected individuals lose their life due to the virus. In this article, we have assumed that the incubation period of the virus is 14 days for patients in India. From the chart presented above, we describe the SIRD modeling of the virus by the time rate of change of the different compartments of the population using coupled ordinary differential equations.

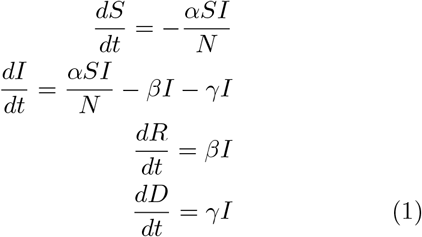

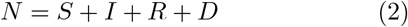

The much talked about *R*_0_ value of a viral epidemic is the number of individuals to whom one infected person can transmit the virus in a day. It can be calculated by using the expression *R*_0_ = *α* × Incubation period (Incubation Period 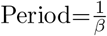, if appropriate unit is used). If *R*_0_ values become less than 1, then we can think that the situation will be under control as the disease now will eventually die down. It is so because the number of people infected per day would become less than the number of people recovering during that period.

We present the plot of the cumulative data of I, R, and D, obtain the parameters, *α, β*, and *γ* by the curves for fixed time intervals with multiple iterations. Later, we solve the mentioned differential equations for the multiple sets of parameters, and their corresponding time intervals, which nearly fits the data. Once the phase-wise plotting is done, we note down two sets of parameters, one corresponding to a no-lockdown/constraint system and another with some constraints. It must also be noted that this analysis assumes the occurrence of both extremities, and the period of the analysis is from fourth march 2020 to 12th May 2020.

### B. Ising model

On the other hand, we use the Ising model [3] to build a quantum circuit to demonstrate the effect of ‘curve flattening’. The Ising model of atomic spin discusses the spin interaction of an individual atom in the lattice with its neighboring lattice points and how the spins behave in the presence of a magnetic field. Each can have an atomic spin of 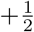 or 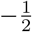. The model describes the spin-spin coupling and the exchange interaction between the lattice points and the associated energy value to them. Interaction matrix depicts the fashion in which two lattice points interact with each other.

Here, Squillante *et al*. [3], attempted to study the COVID-19 spread by comparing it with the Ising model, with the analogy of each atom as an individual. The spin of each lattice point (individual) describes whether the person is infected with the virus or not. A spin of 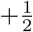 (p=probability of getting infected [success]) depicts that the person is infected with the virus and the spin of 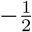 (q=1-p, probability of not being infected [failure]) indicates otherwise, while considering the total population = N. Hence, just like the Ising model, here also the spins (infected or susceptible individuals) interact with each other. The effect of magnetic field has not been considered since it doesn’t carry in relevance in the analogy to the viral spread.

Since, in the case of viral epidemics, the infection is spread through ‘contact’ between individuals [13, 14], we assume in a population of N, out of which ‘S’ are susceptible ‘r’ people are infected. Here, we consider when two infected people interact, the net interaction is 0 or no effect. So a general probability distribution can be obtained from the Bernoulli’s equation as follows-

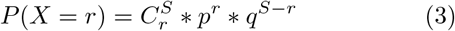

From the previous SIRD model, we get a reproduction rate 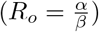 which also tells about the average number of secondary cases arising from primary cases in an entirely susceptible population, so we can consider 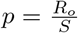 or *pS* = *R*_*o*_. The resultant equation turns out to be

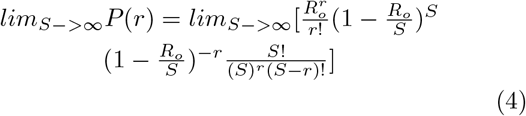

On using Stirling approximation **??** formula and further solving (4) we get a Poisson distribution (for further details, refer Appendix VI),

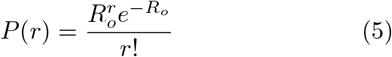

As analysed, new cases per day rise nearly exponentially to maximum value and then decrease, which shows the trend of a peak function, from which the equation could be derived (17). We can derive its time evolution factor with the probability of cases by implementing Hamiltonian operator of its function dependent on time. The spin systems interact with each other by the exchange interaction. It is defined by a Hamiltonian operator as-

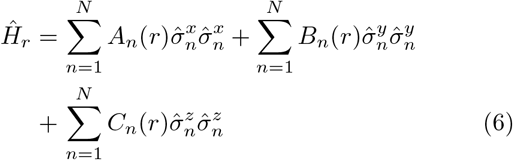

where, *A*_*n*_, *B*_*n*_ and *C*_*n*_ are the probability functions. The strength of the exchange interaction decreases as the distance between these particles increases (which can also deduce that if people maintain distance, the spread of infection may decrease). In the neighboring region, we can almost assume that the interaction of one particle is almost similar to all of the neighbors, and the interaction is followed in all three directions, and thus the exchange interaction takes place; the stronger the exchange interaction more will be its infection rate.

Hence to find its co-relation with the time evolution operator and deriving it into quantum circuits here, for n=1, we get a more straightforward form i.e.

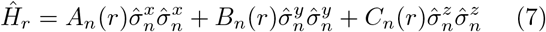

We apply the time evolution unitary operator on our Hamiltonian operator

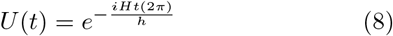

by taking 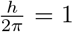 and putting the Hamiltonian of (7) in unitary operator of (8) we deduce.

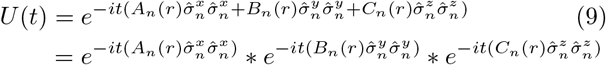

where, 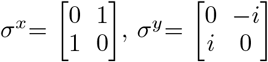 and 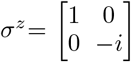

Dividing the equation (10) into 3 parts as –

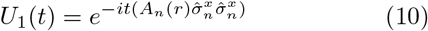

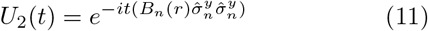

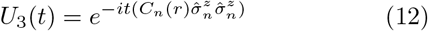

On solving the equation (10)

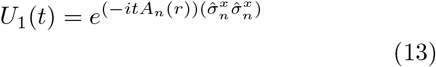

which follows the condition of

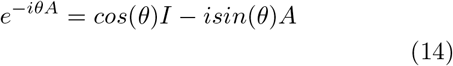

where I is the identity matrix and A is of same order of the identity matrix.

Here, A=*σ*_*x*_ ⊗*σ*_*x*_, which follow the identity rule and has a order 4 and we *break* down it to

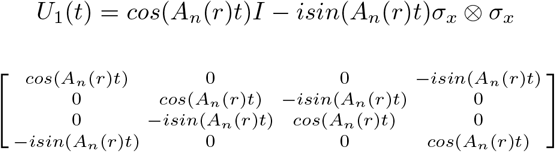

Hence, solving the matrix and comparing with the U3 matrix we derive a following circuit 1:

Similarly, working for the equation 11 we get a reduced matrix format

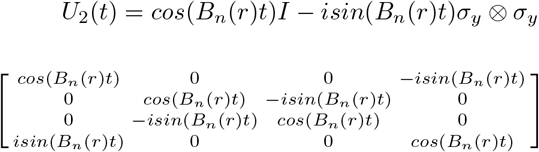

Where the equivalent matrix is reduced in the form of circuit as, 2

Similarly, working for the equation 12 we get a reduced matrix format

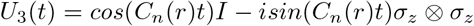

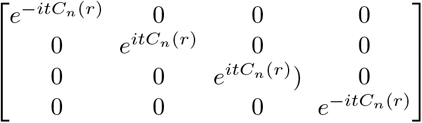

Where the equivalent matrix is reduced in the form of circuit as, 3

Combining these 3 circuits, we get an equivalent circuit as in Ref. 4,

Finding the average value helps us find the Energy eigenstate of the operator, which we can relate to the number of infected people at a particular time.

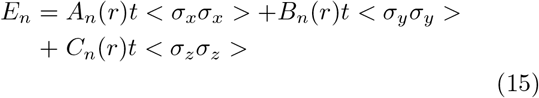

where, *<*> = shows average value and

<*σ*_*x*_*σ*_*x*_ >= *P*_00_ − *P*_01_ − *P*_10_ − *P*_11_

which is determined by putting 2 Hadamard gates at the end of the equivalent circuit, which is useful for moving information between the x and z bases, which is shown in the following diagram (5).

Similarly, <*σ*_*y*_*σ*_*y*_ > can be represented by putting a inverse S gate followed by a H gate in each qubit as it moves information from y to z bases, which is shown in the following diagram (6).

And for <*σ*_*z*_*σ*_*z*_ >, we only measure the qubits in zz basis, which is shown in the following diagram (7).

*A*_*n*_(*r*), *B*_*n*_(*r*) and *C*_*n*_(*r*), all three functions are taken to be P(r) which we have derived in the above. Hence the Hamiltonian, whose time evolution we study for COVID-19 spread now turns-

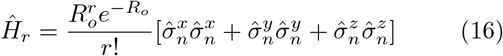

## III. IMPLEMENTATION AND RESULTS

### A. SIRD model

The first Covid-19 case was reported in India on 30th January 2020. Considering it as Day 1, on 4th March (the 33rd day), the number of cases jumped suddenly from 5 to 28. So, in this study, we start analyzing the data from March 4th onwards. For solving the differential equations, we use the Odeint module from scipy, which uses the LSODA algorithm to solve the system of coupled ODEs. As Rajesh *et al*. [10] pointed out in their SIRD model prediction of Covid-19 in India, that there is no reported error in the database, so we cannot use reduced chi-sq fitting for the above data. So, we employed the approach mentioned to carry out our analysis i.e., the method of eye-approximation for the best fit.

As per this fitting, in Fig. 8, the peak of the infection curve suggests that a maximum of 10^9^ people will get infected for a particular set of parameters. However, data plotting suggests a change in the concavity of the curve it follows periodically, hence in its slope. Thus taking a single fitting for the whole period might not be a well scientific method. Hence, we tried to break the time interval into smaller legs (here ten days) as per Table I and record the values of the parameters (Table II) in the respective leg. The simulated curves are thus made to nearly fit the data points in Fig. 9.

**TABLE I:**
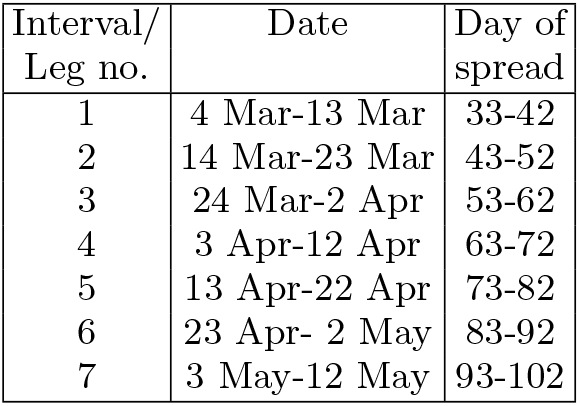
Distribution into various legs

**TABLE II:** Values of *α*(t) and *β*(t) for various intervals for making the curve fit with the respective data sets.

| Interval/<br>Leg no. | $\alpha$ | $\beta$ |
| --- | --- | --- |
| 1 | 0.127 | 0.002 |
| 2 | 0.167 | 0.010 |
| 3 | 0.165 | 0.014 |
| 4 | 0.146 | 0.014 |
| 5 | 0.101 | 0.017 |
| 6 | 0.0825 | 0.020 |
| 7 | 0.091 | 0.024 |

**FIG. 1:**
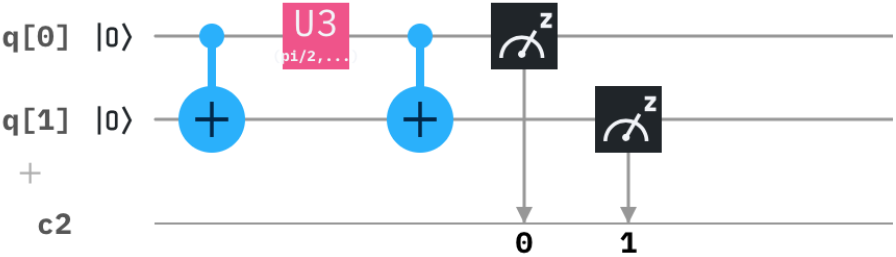
The derived circuit has a combination of a CNOT gate, U3 gate and a CNOT gate, where *θ* = 2*A*_*n*_ (*r*)*t*, *ϕ* = - *π*/2 and λ = *π*/2.

**FIG. 2:**
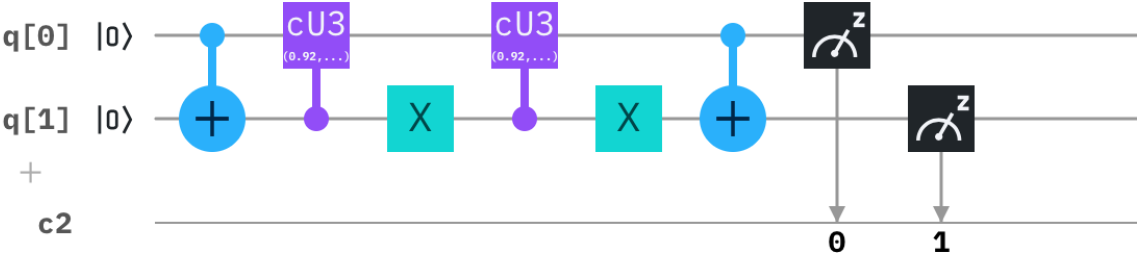
The derived circuit has a combination of a CNOT gate, control U3 gate, Anti-control U3 gate and a CNOT gate, where *θ*_1_ = 2*B*_*n*_(*r*)*t, ϕ*_1_ = −*π/*2 and *λ*_1_ = *π/*2 and *θ*_2_ = 2*B*_*n*_(*r*)*t, ϕ*_2_ = *π/*2 and *λ*_2_ = −*π/*2.

**FIG. 3:**
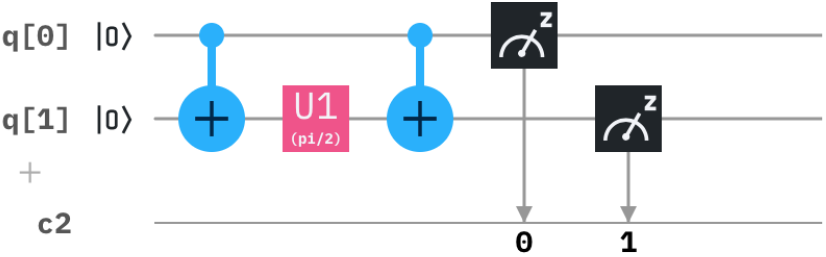
The derived circuit has a combination of a CNOT gate, U1 gate and a CNOT gate, where *θ* = 2*C*_*n*_(*r*)*t*.

**FIG. 4:**
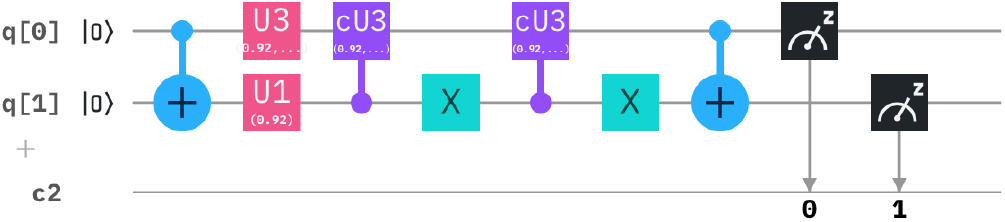
The equivalent circuit describes the Hamiltonian operator 7 in a N=2 state.

**FIG. 5:**
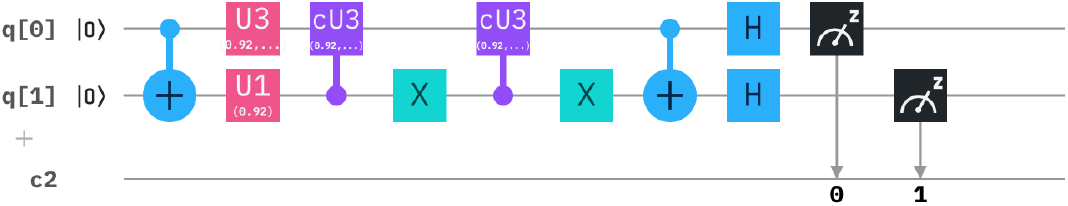
Measurement bases for the calculation of average values of *< σ*_*x*_*σ*_*x*_ *>* for N=2 in circuit.

**FIG. 6:**
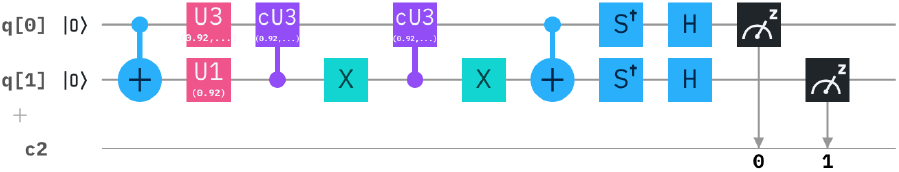
Measurement bases for the calculation of average values of *< σ*_*y*_*σ*_*y*_ *>* for N=2 in circuit.

**FIG. 7:**
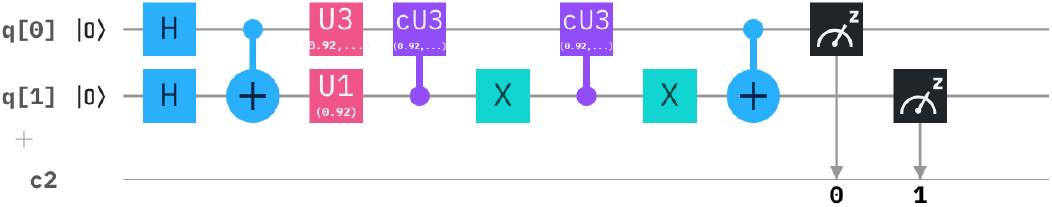
Measurement bases for the calculation of average values of *< σ*_*z*_*σ*_*z*_ *>* for N=2 in circuit.

**FIG. 8:**
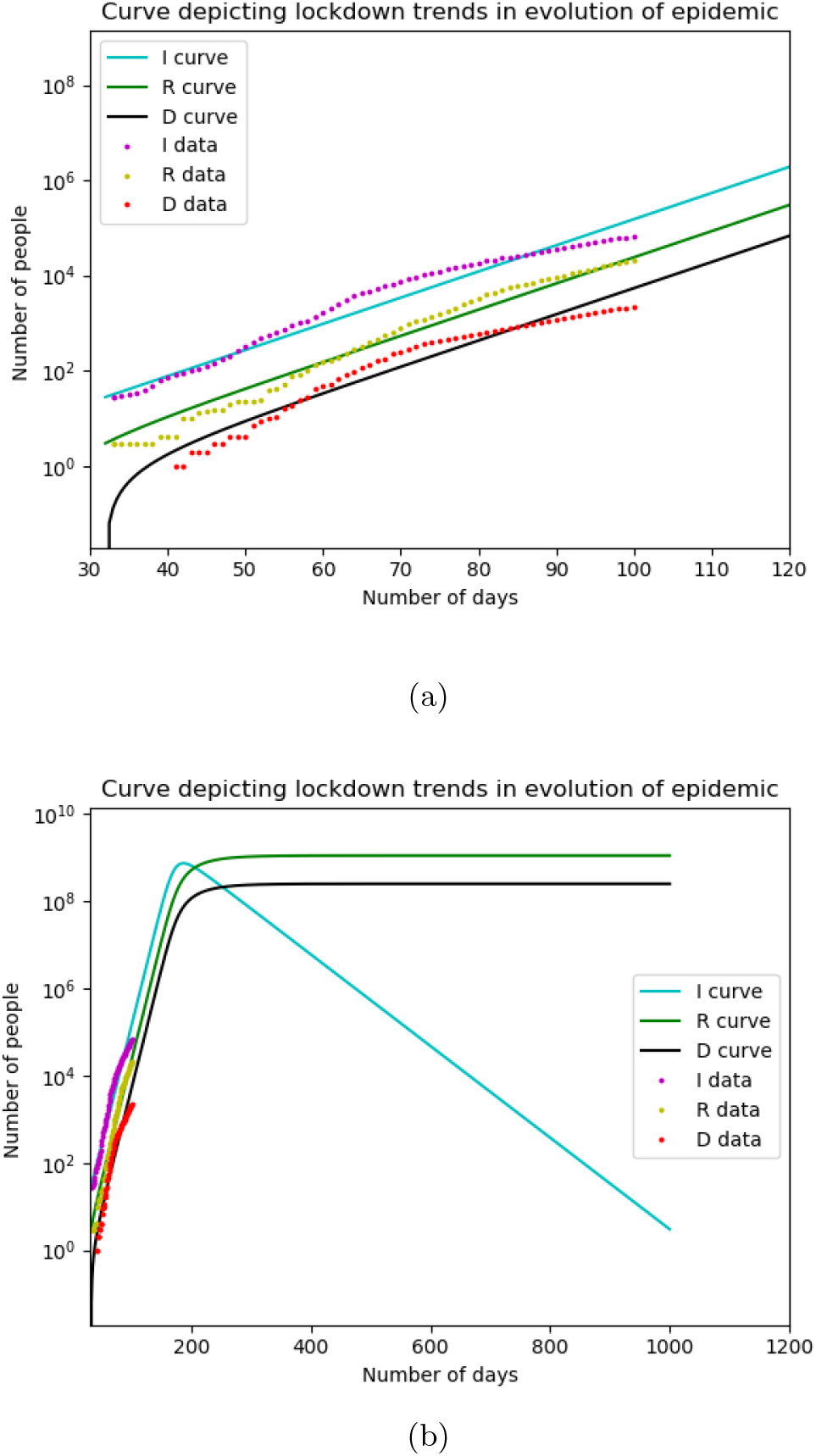
Simulating the prediction curves by using the best fit over the entire range of data. Fig. (a) depicts the data points and respective curves with *α* = 0.144, *β* = 0.021 and *γ*=0.0041 (b) represents the same curve with the time axis extended upto 1000 days, thus giving a trend of the infection.

**FIG. 9:**
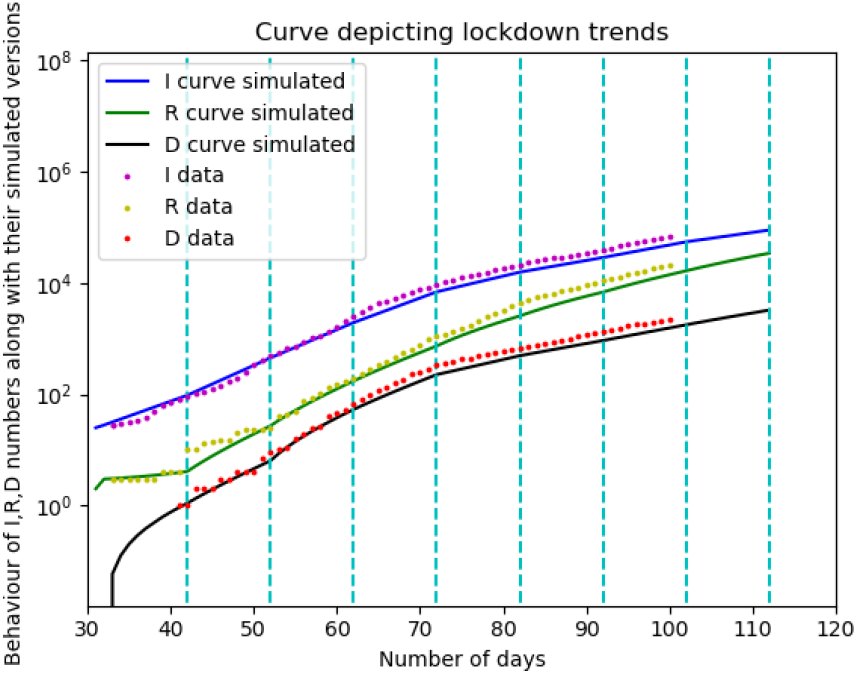
Simulating the prediction curves by using the best fit for every single interval obtained by dividing the time axis by the dashed cyan lines.

By obtaining values of parameters, we proceed to make the curves for infection, recovery, and deaths that approximately match the data points available. Furthermore, thus we obtain alpha and beta as for each period i.e., *α*(t) and *β*(t). Here, our only concern remains *α*(t), and how it evolves in time as it is dependent on interactions between components of the population that’s being curbed and regulated during a lockdown [4]. *β*(t) and, on the other hand, is dependent on various other factors like the efficacy of treatment, the capacity of the healthcare system, and demographics of the individual population, which is not the point of discussion in this paper, hence not given much importance. Also, changes in their values are in the order of 10^−3^ if any were made during the fitting and duly presented.

In India, many cases start exhibiting the symptoms after completing their 14-day quarantine. Hence, for safety purposes, many state governments have extended their quarantine period to 28 days. So it might be unsafe to assume any incubation period of fewer than 14 days. Hence we consider it to be exactly 14 days. Moreover, that makes the R0 value for the 3rd leg (March 24-April 2) 2.31, which falls to 1.274 by the end of the 7th leg. Hence, the lockdown has been able to contain the spread to some extent, but it should continue until it reaches a value closer to 1.

If we go on plotting the evolution of *α*(*t*) from the above data, we can see that a lockdown can successfully reduce the infection rate. To know the future of *α* under the lockdown, we consider the rate of infections of the several smaller periods under lockdown while excluding the first 2 data points for day 33-42 and 43-52 (since the lockdown nearly began on the day 53 of the arrival of the virus in India) and do an exponential decay fit. We later go on extrapolating the curve to know the estimated alpha values in the few next legs. As shown in figure 10, we later use a straight-line fitting to extrapolate beta for subsequent legs. Here, we can interpret that the recovery rates have increased in a pretty uniform manner. It can be attributed to various aspects, such as improvement in the healthcare scenario, change in the demography of the infected individuals, or a boost in the quality and quantity of available treatment methods. Further studies and accurate models might be able to justify this trend (Fig. 10).

**FIG. 10:**
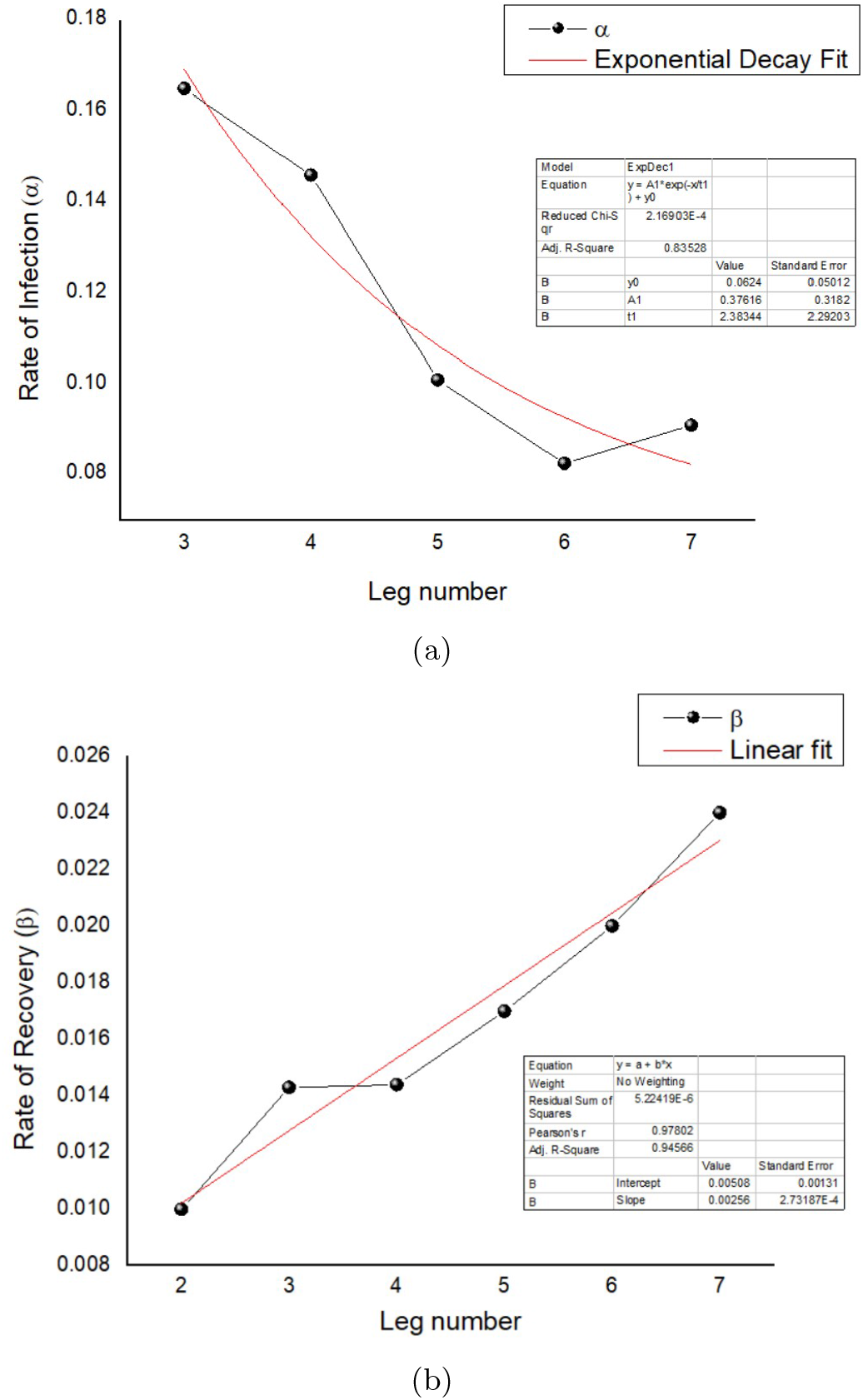
Curve fitting for studying the evolution of *α* and *β* over time. Fig (a) *α*(t) being fit to an exponential decay function *y* = 0.063 + 0.376 · *e*^−*x/t*^ (b) *β*(*t*) being fit yo a straight line *y* = 0.00256*x* + 0.00508.

We use the obtained values of *α* and *β* for the next legs and simulate the SIRD curve and present it cumulatively. We have tried to analyze how the future numbers might be if the lockdown continues as it is until the disease is entirely null. After 3-4 legs further, the change in *α* would have started becoming negligible (and R0 value slowly tending to 1). The recovery rate also would have become nearly constant, we have taken the values of the parameters at the end of the leg of period 132-142 to be constant throughout, for the time to come. We extrapolated the curve to obtain a trend (Fig. 11).

**FIG. 11:**
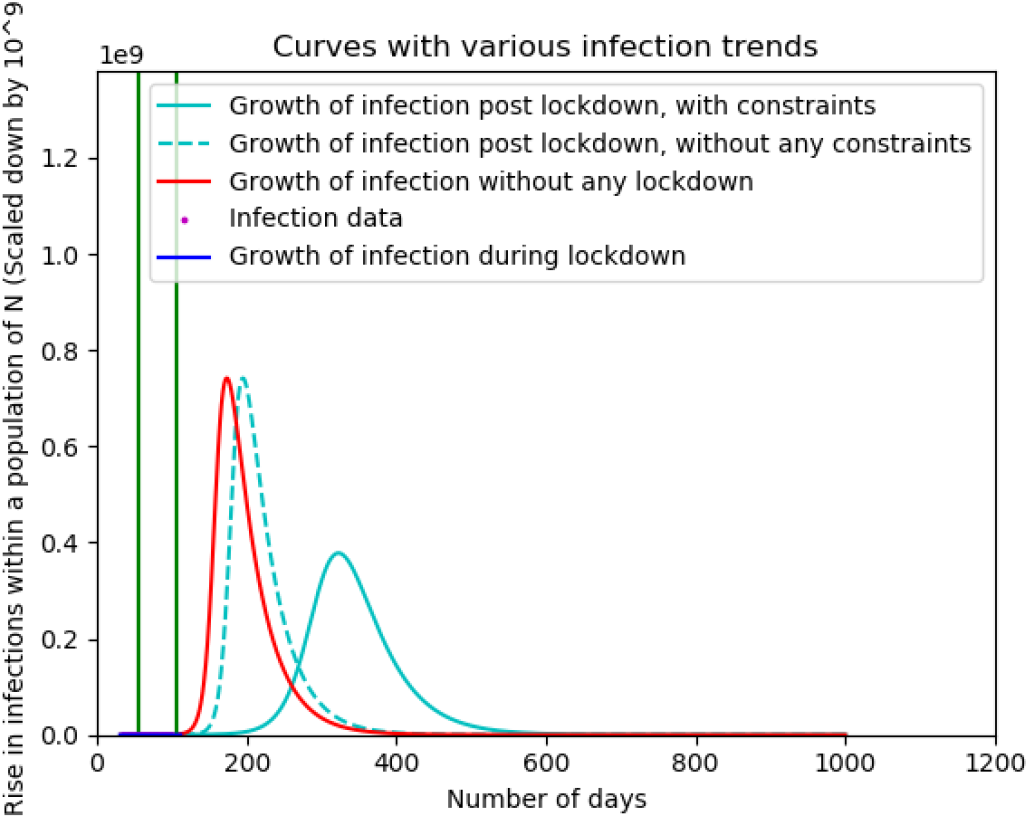
Analysis of number of infections upon various scenarios of lockdown. The time axis is segmented by using green lines, which signify the start and (Probable) end of lockdown. It can be observed that the number of cases at the peak almost halves with a distinct shift in the abscissa of peak, when the lockdown continues. When lockdown is removed all of a sudden (and assumed that the *α*= 0.165, just like the value before lockdown), there is less decline in the number of cases at the peak with a subtle shift in the abscissa of peak.

Parallelly, we have studied the effect of the current lockdown being lifted. If the lockdown is lifted on May 17, we had assumed the other extreme that the infection rate might well reach the value it had before the imposition, but certainly not anything greater than that and plotted the curves.

### B. Ising model

From Equation (16) from the previous section, we have a Hamiltonian for the COVID-19 spread when compared to the Ising model. We were able to find a function which, when fed into the quantum circuit, qualitatively depicts the shape of the curve for the infected individuals, as we also obtained in the SIRD modeling.

While trying out various functions, we finally found that a Poisson probability distribution function gives a considerable resemblance to an actual graph and also the type obtained from SIRD modelling (Fig. 12). In the function, the parameter *R*_0_ signifies the ratio of *α* to that of *β*. We can observe that when *R*_0_ is decreased while keeping other variables and parameters constant, we can observe that the peak of the curve shifts forward in the time axis with a reduction in its height and making it broader.

**FIG. 12:**
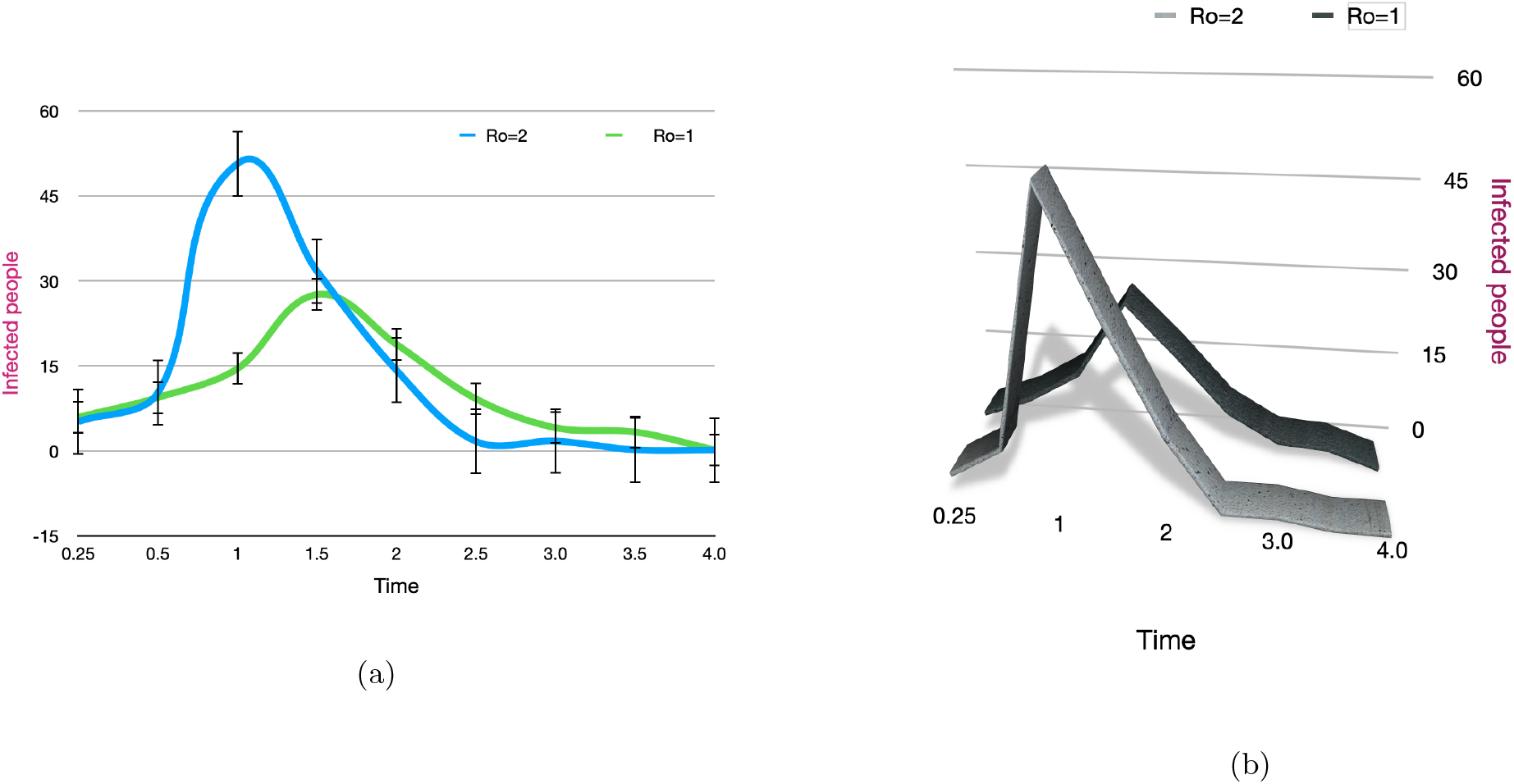
Used Hamiltonian being simulated using the IBM Quantum Experience, demonstrating the phenomena of curve flattening. Fig (a) A 2-D graph depicting the shift of the peak along X-axis when *R*_0_ is decreased (b) A 3-D curve showing the decline in the height of the peak when *R*_0_ is decreased.

From these, we can easily derive that a decline in the rate of infection (*α*) will infect lesser people than before, thus flattening the curve. Also, an increase in the rate of recovery (*β*) will imply an improvement in healthcare facilities, changing demographics of the infected patients, etc. and show a change in the height and position of the peak. Using proper parameters and constants, this can be a novel way to simulate approximate curves for given values. An analysis of epidemic spreads can be carried out with the inclusion of more and more factors that otherwise gets difficult for classical computers.

## IV. INTERPRETATION AND DRAWBACKS

The three graphs, with I, R, D, show two trends post lockdown-one set of parameters without any distancing (constraints), and their respective carrier limits, and time is taken to attain them. The latter case depicts another set of parameters that have a few constraints imposed on them, as in the last phase of lockdown. So latter case parameters postponed the attainment of carrier limits and are not as rapidly growing as the former case. Also, the no lockdown case is started from the day of data collection, along with lockdown trends and post lockdown trends growing by two parameters to give an effective picture of what would have been the picture without a lockdown and how it would evolve once a lockdown is lifted off considering the extreme case scenario too. It is evident that the lockdown shouldn’t be lifted all of a sudden, as it will lead to a massive upsurge in the number of cases (nearly same as that of no-lockdown case) with a very narrow delaying of the peak.

The SIRD model discussed here has its own set of limitations pertaining to the assumptions made and the methods adopted while formulating it. The first limitation of this analysis is the use of the slope-estimation of data plotted curves to find the simulated curve in the lockdown period while obtaining a periodic split up of parameters. This might account for errors in the analysis. The assumptions like not considering birth and mortality rates, the permanent immunization of a recovered patient, unavoidable migration and interactions, etc. make the model lose its precision. Here, the susceptible are considered to be the entire population except the infected, recovered, and deceased individuals, initially at a time t. No considerations have been made to distinguish the exposed individuals out of the susceptible and model them accordingly. This might be a significant drawback of the model. The trend in recovery and its rate also is incomprehensible due to the lack of data on the demographics of the recovered patients in terms of their age, sex, etc. In this model, the number of tests performed and their rate has not been considered, but they might play a significant role in reporting the cases, hence the *α*(t).

Our N (the total population) has been assumed constant, but immigration, new birth, and deaths might vary the N, thus making us compromise on its accuracy. More efficient models like the SEIRD (Susceptible-Exposed-Infected-Recovered-Deceased), and higher compartmental models can be implemented for better prediction and analysis.

The quantum circuit design only gives the qualitative shape of a ‘usual’ curve of an epidemic infection spread with the proper use of the parameters and some constants, this can be used to simulate the curves for respective regions. Adding various complexities, out of which some are discussed in the above, is a matter of time. With better and smarter use of quantum gates, efficient circuits can be made to minimize the space and time complexities. Here, we have assumed only two parameters, the rate of infection and the rate of recovery. This can be intricately designed to accommodate various other detailed parameters that encompass factors like the number of tests, the age group of infected individuals, government measures in curbing the epidemic, etc. Moreover, this can give us a more accurate simulation of the epidemics as significant as COVID-19.

## V. CONCLUSION

COVID-19 has turned out to be a global crisis, affecting all the countries. The current model study report hints at a frightening upsurge of the viral epidemic in the times to come. Measures like quarantine and lockdown have been successful enough to reduce their impact, yet a lot needs to be taken care of. However, the current prolonged lockdown has started worrying national as well as global economies, pushing them into a tremendous crisis; hence the lockdown cannot be sustained forever. But as per our analysis, the lifting of lockdown shouldn’t be all of a sudden, and be more gradual in the approach. We must start practicing the concept of social distancing and personal hygiene to keep the viral spread at bay. In desperate times like these, researchers of all fields must come together and contribute towards finding more information regarding the virus through experiments and data analysis. This shall let us be more aware and help us in tackling the risk. We sincerely hope that, just like previous global pandemics, we can pass through this with the advent of science and technology.

## Data Availability

All the data have been imported from COVID-19 India Tracker, it is a crowd-sourced database.

https://www.covid19india.org/

## ACKNOWLEDGMENTS

S.P., A.P., P.S. and K.S. would like to thank Bikash’s Quantum (OPC) Pvt. Ltd. for providing hospitality during the course of this project. A.P. would like to thank Dr. Victor Roy, SPS, NISER for clearing some of the doubts and sharing valuable resources in the initial phase of the project. B.K.B. acknowledges the prestigious Prime Minister’s Research Fellowship awarded by DST, Govt. India. The authors acknowledge the support of IBM Quantum Experience for producing the basic circuits. The views expressed are those of the authors and do not reflect the official policy of IBM or IBM Quantum Experience team.

## VI. APPENDIX

p is the probability of getting infected.

q is the probability of not getting infected,i.e.,q=1-p.

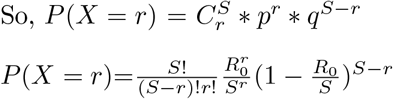

Assuming the susceptible to be ifinite,R0 is a constant.

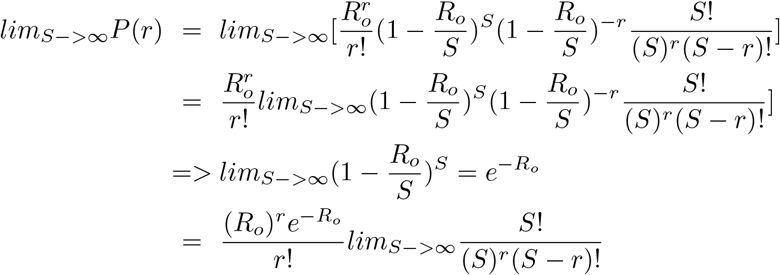

*Using* : *Stirling′s* − *approximation* − *formula*

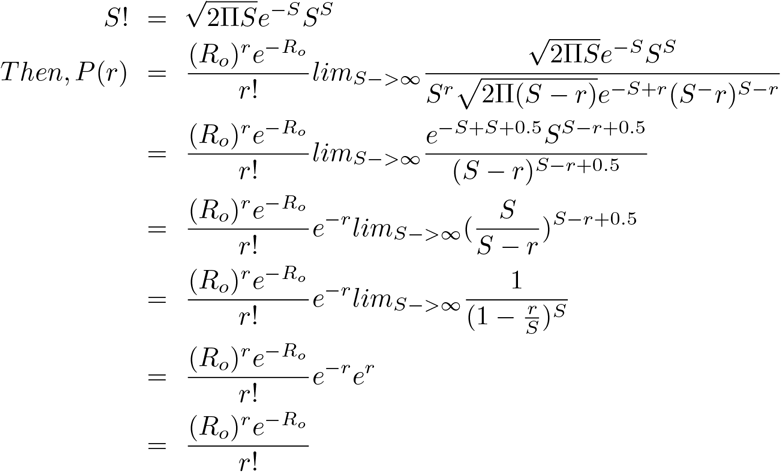

